# Human Mobility and Infection from Covid-19 in the Osaka Metropolitan Area

**DOI:** 10.1101/2022.05.12.22274931

**Authors:** Haruka Kato, Atsushi Takizawa

## Abstract

Controlling human mobility is thought to be an effective measure to prevent the spread of the COVID-19 pandemic. This study aims to clarify the human mobility types that impacted the number of COVID-19 cases during the medium-term COVID-19 pandemic in the Osaka metropolitan area. The method used in this study was analysis of the statistical relationship between human mobility changes and the total number of COVID-19 cases after two weeks. In conclusion, the results indicate that it is essential to control the human mobility of groceries/pharmacies to less than 0% and that of parks to more than -20%. The most significant finding for urban sustainability is that urban transit was not found to be a source of infection. Hence governments in cities around the world may be able to encourage communities to return to transit mobility, if they are able to follow the kind of hygiene processes conducted in Osaka.

## 1. Introduction

### 1.1. Background

The COVID-19 pandemic was reported to positively impact urban sustainability in the short term [1]. However, the pandemic might not continue to impact urban sustainability positively even in the medium term. For example, several lockdowns caused changes in mobility, especially in public transportation, because of concerns about COVID-19 infection [2]. However, the modal shift from public transportations to cars will adversely effect on reduction of carbon dioxide emissions [3]. Controlling human mobility is thought to be an effective nonpharmaceutical intervention to prevent the spread of the COVID-19 pandemic worldwide, for example, in the United States, the EU, and China [4, 5, 6]. To control human mobility, states of emergency have been declared many times during the medium term COVID-19 pandemic. For example, the Japanese government declared states of emergency several times in Osaka Prefecture [7]. During these emergency declaration periods, the Subcommittee on Novel Coronavirus Disease Control in Japan requested that residents reduce their human mobility by 50% [8]. During the first emergency declaration, this requirement reduced the home range in suburban cities of the Osaka metropolitan area by 50% due to residents changing their transportation methods to walking and cycling [9,10,11]. During the same first emergency declaration, a decrease in human mobility was also reported in Tokyo [12]. However, some studies have suggested that the effects of controlling human mobility differed in the early term and medium term [13,14,15].

This study investigates the following research question: Where should we control human mobility to reduce the number of COVID-19 cases? The effectiveness of controlling human mobility has been reported through the implementation of school and company closures [9,10,11]. In addition, in Japan, many prefectural governments requested that dining and drinking establishments close by 8:00 p.m. and refrain from serving alcohol [7]. However, to prevent the spread of the COVID-19 pandemic, it is necessary to consider restricting places that have not previously been considered. In addition, to maintain socioeconomic activities, it is essential to consider ending restrictions in places where infections are less likely to occur. Therefore, the type of human mobility contributes to how policymakers develop policies to control the spread of infection for urban sustainability.

### 1.2. Purpose

This study aims to clarify the human mobility types that impacted the number of COVID-19 cases during the medium-term COVID-19 pandemic in the Osaka metropolitan area. The method used in this study was analysis of the statistical relationship between human mobility changes and the total number of COVID-19 cases after two weeks. The human mobility types are divided into six categories using Google Community Mobility Reports data: retail and recreation (retail/recreation), groceries and pharmacies (groceries/pharmacies), parks, transit stations, workplaces, and residential areas [16]. These human mobility types are analyzed based on the relative changes in the number of visitors to the six types of places. Random forest analysis is applied to determine this statistical relationship.

The Osaka metropolitan area from March 1, 2020, to September 30, 2021, was selected as a case study. During the medium term, the Osaka metropolitan area experienced five waves of increasing and decreasing numbers of COVID-19 cases. During this period, vaccination started for healthcare workers in February 2021 and for older people and adults in April 2021. As a result, the fourth state of emergency was lifted in the Osaka metropolitan area on September 30, 2021. In this study, the Osaka metropolitan area consists of Osaka Prefecture, Kyoto Prefecture, and Hyogo Prefecture, as shown in Figure 1. Unlike other metropolitan areas, the Osaka metropolitan area has three central areas: Umeda in Osaka Prefecture, Karasuma in Kyoto Prefecture, and Kobe in Hyogo Prefecture. Because multiple railways and highways connect these areas, the number of infections tended to increase speedily. In the early term COVID-19 pandemic, it was found that city size correlated with COVID-19 cases [17]. Therefore, COVID-19 countermeasures were often discussed and collaboratively implemented by the governors of the three prefectures. For example, the three governors together requested that the Japanese national government declare a state of emergency. In addition, the emergency declaration periods have been the same for all three prefectures. Therefore, it is reasonable to analyze these three prefectures together as the Osaka metropolitan area.

**Figure 1.**
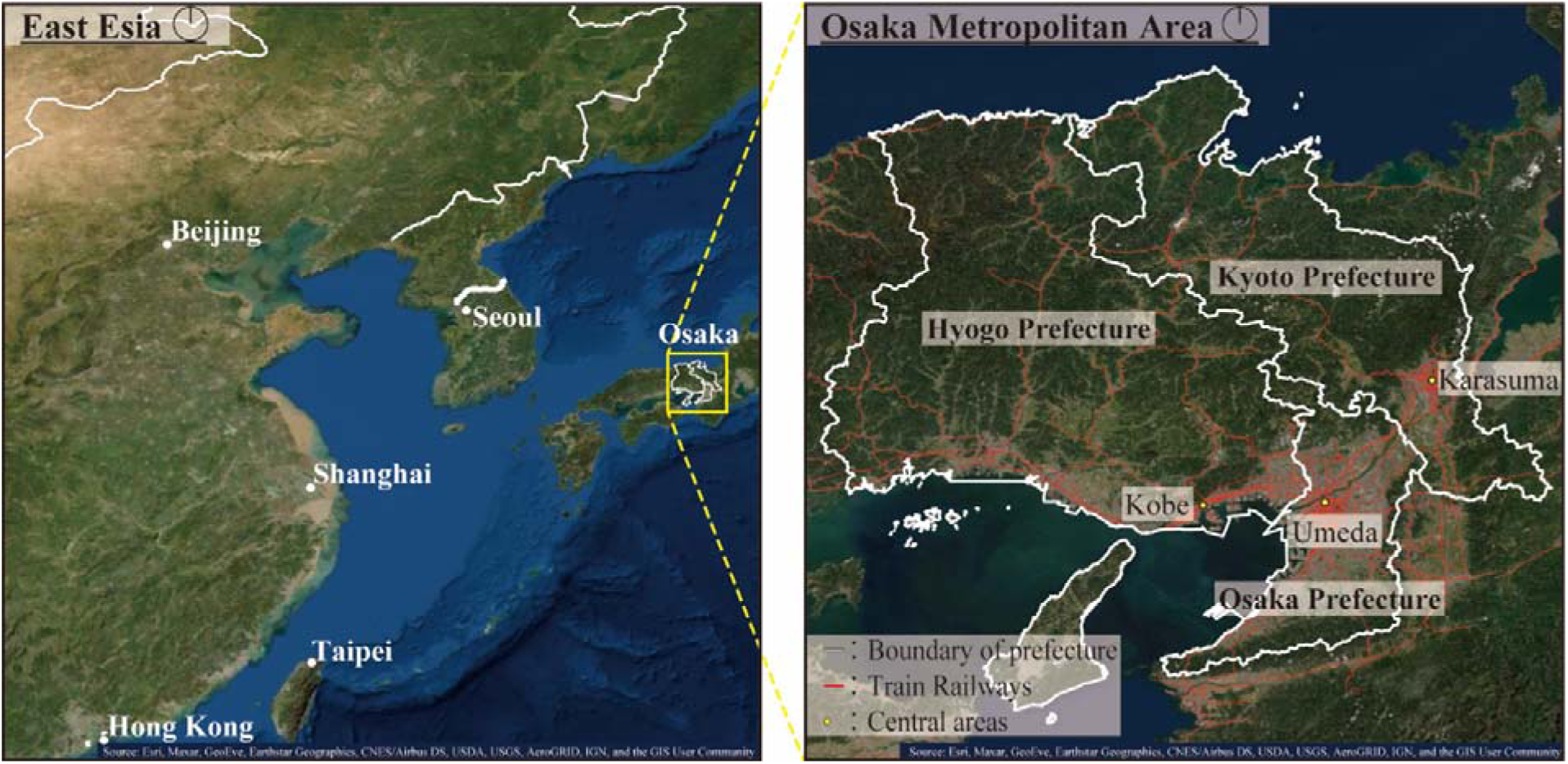
Map of the Osaka metropolitan area in Japan. In this study, the Osaka metropolitan area consisted of Osaka, Kyoto, and Hyogo Prefectures. The Osaka metropolitan area has three central areas with multiple railways.

### 1.3 Literature Review

Many studies on human mobility have used mobile phone data to predict the number of people infected with SARS-CoV-2. Restaurants, fitness centers, cafes, bars, and hotels have been found to be high-risk areas for infection [18]. In Japan, the human mobility of downtown nightlife was found to be higher risk than that of residences or workplaces [19]. In addition, prefectural governments have often considered COVID-19 countermeasures with reference to the human mobility of transit station [20]. For example, the Osaka Prefectural Government alerted people when the human mobility increased at the terminal stations [20]. Similar to this study, random forest analysis using Google Community Mobility Reports data indicated that the important mobility areas were retail/recreation, groceries/pharmacies, and transit stations in the case of the EU from March to April 2020 [21]. In addition, in Germany from February to July 2020, the factors related to the number of cases were increasing human mobility of groceries/pharmacies and decreasing mobility of workplaces and retail/recreation [22]. Regarding individual human mobility, human mobility was reduced for workplaces and transport during the pandemic in Portugal [23]. The results indicated that workplace closure was nearly as effective as the stay-at-home order in the social distancing policy [24]. Japan had one of the greatest declines in human mobility worldwide [25]. In addition to mobility of workplaces, human mobility of parks has received attention. The number of urban park visitors increased worldwide during the pandemic [26].

Based on previous studies, the novelty of this study is its clarification of human mobility types during the medium-term COVID-19 pandemic. In Japan, the state of emergency was called a “soft lockdown” because the Japanese government did not restrict the activities of individuals [27]. For example, the Osaka Prefectural Government demanded that railroad companies conduct temperature checks at major terminal stations and move up the last train departure time [28]. Besides, based on the demand by the prefectural governments, railroad companies decided to remain the number of daytime train departures the same as before the pandemic, even if the number of passengers decreased significantly [29]. Some railroad companies in Osaka also provided incentives to those who took the train when the number of passengers was less [30]. Those hygiene processes reduced the density of human mobility at the transit station. Therefore, most citizens could go out at least occasionally, even under the state of emergency declaration. Therefore, there might be diversity in the relationship between human mobility types and the number of COVID-19 cases. The results will help policymakers plan effective human mobility control for urban sustainability. However, it is difficult to obtain highly accurate results for the medium-term COVID-19 pandemic [13,14,15]. Previous studies have analyzed the relationship between the number of COVID-19 cases and human mobility using the logistic growth model [31], partial differential equation [32], and neural network [33]. Referring to the analysis of Delen *et al*. [22], this study uses random forest analysis to obtain highly accurate results.

## 2. Results

### 2.1 Changes in Human Mobility

#### 2.1.1 Human Mobility Changes from March 2020 to September 2021

Figure 2 shows daily changes in human mobility in the Osaka, Kyoto, and Hyogo Prefectures for the medium-term COVID-19 pandemic. Figure 2 shows the spline curve and the confidence interval. The smoothing parameter of the spline curve λ was set to 0.001. Additionally, Figure 2 indicates the emergency declaration period. The results show similar changes in the Osaka, Kyoto, and Hyogo Prefectures.

**Figure 2.**
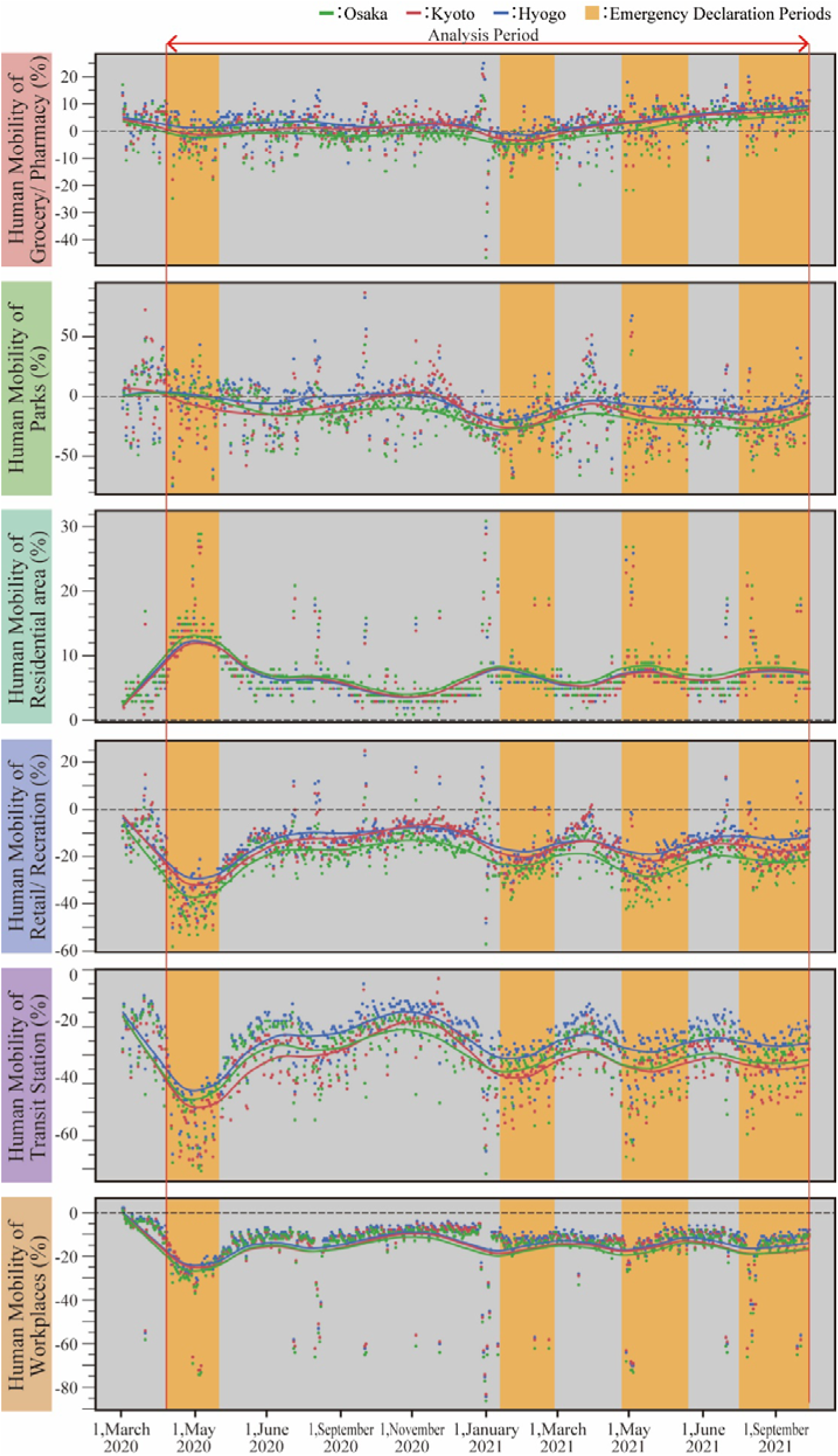
Human mobility changes from March 2020 to September 2021. Human mobility types are retail/recreation, groceries/pharmacies, parks, transit stations, workplaces, and residential areas. Green points and line are the data of Osaka Prefecture, red points and line are the data of Kyoto Prefecture, and blue points and line are the data of Hyogo Prefecture.

Figure 2 shows that human mobility varies according to the six types. After March 2020, all types of human mobility except human mobility of residential are decreased. This finding suggests that more people stayed at home, even without the stay-at-home order. When human mobility of residential are increased, other types of human mobility decreased. Due to the emergency declaration, human mobility decreased in transit stations and retail/recreation. In addition, the human mobility of workplaces declined sharply during holiday periods, such as summer vacations and the new year holiday. The human mobility of groceries/pharmacies remained at approximately 0%, although it changed slightly during the emergency declaration. The human mobility of parks increased during the first emergency declaration but then began to decrease.

### 2.2 Change in the Number of COVID-19 Cases

Figure 3 shows the daily change in the number of people infected with SARS-CoV-2 in the Osaka, Kyoto, and Hyogo Prefectures. Figure 3 shows the spline curve and the confidence interval. The smoothing parameter of the spline curve λ was set to 0.001. Additionally, Figure 3 indicates the emergency declaration period.

**Figure 3.**
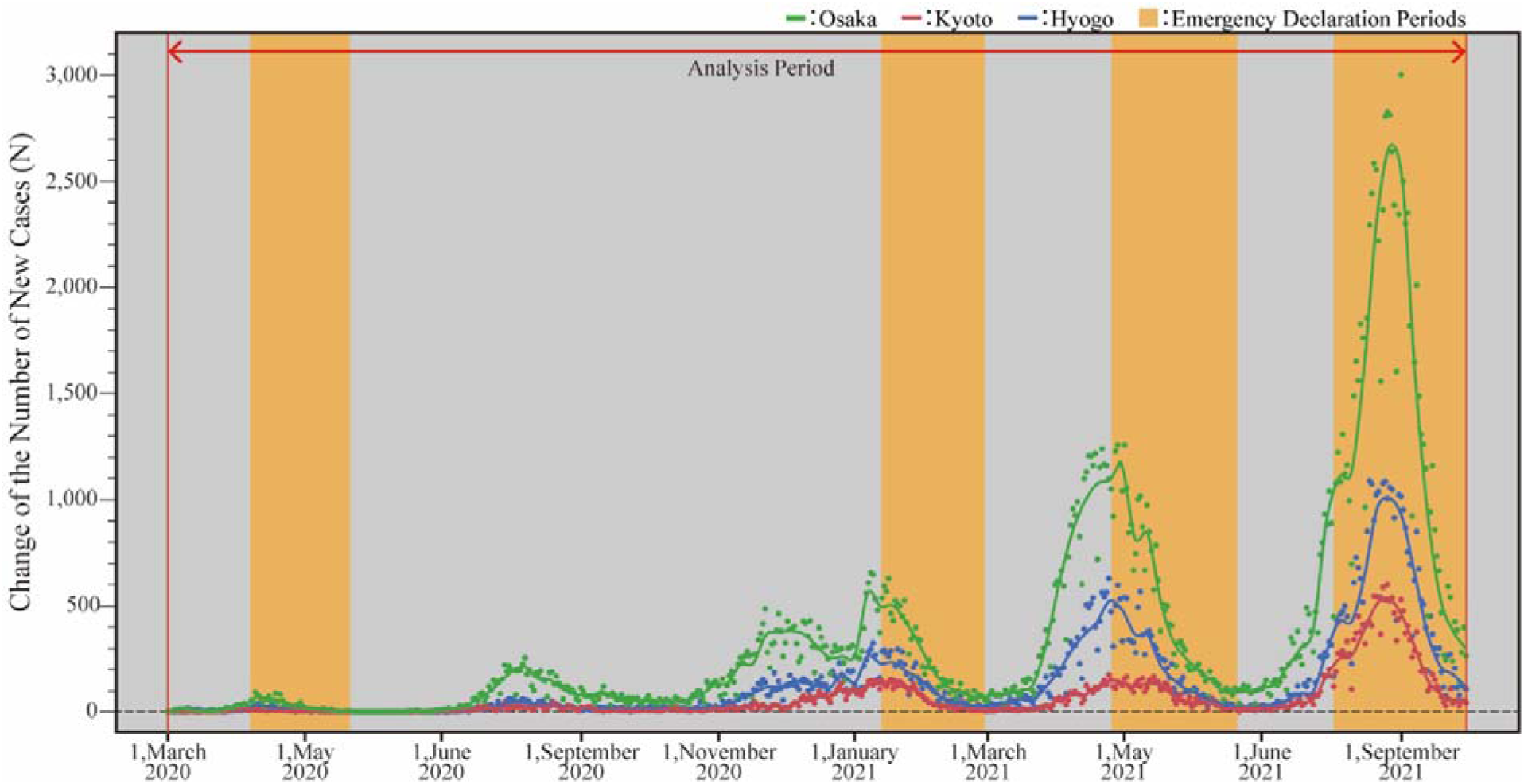
Change in the number of COVID-19 cases. Green points and line are the data of Osaka Prefecture, red points and line are the data of Kyoto Prefecture, and blue points and line are the data of Hyogo Prefecture.

Figure 3 shows that the Osaka, Kyoto, and Hyogo Prefectures experienced five waves of increases and decreases in COVID-19 cases between February 2020 and December 2021. The first wave was from April to May 2020, the second from July to September 2020, the third from December 2020 to February 2021, the fourth from March to June 2021, and the fifth from July to September 2021. The number of infections gradually increased from the first to the fourth wave. States of emergency were declared during the first, third, fourth, and fifth waves. The declaration of a state of emergency effectively reduced the number of COVID-19 cases.

### 2.3 Human Mobility Types That Impact the Number of COVID-19 Cases

Table 1 and Figure 4 show the human mobility types that impacted the total number of COVID-19 cases after two weeks in the Osaka, Kyoto, and Hyogo Prefectures. The statistical analysis was the random forest method. Table 1 shows the main effect and the total effect for each prefecture. Figure 4 shows the variable importance in the Osaka, Kyoto, and Hyogo Prefectures. As shown in Table 1, the R^2^ scores of all models were over 0.7, indicating good accuracy. The results are discussed separately by prefecture.

**Table 1.**
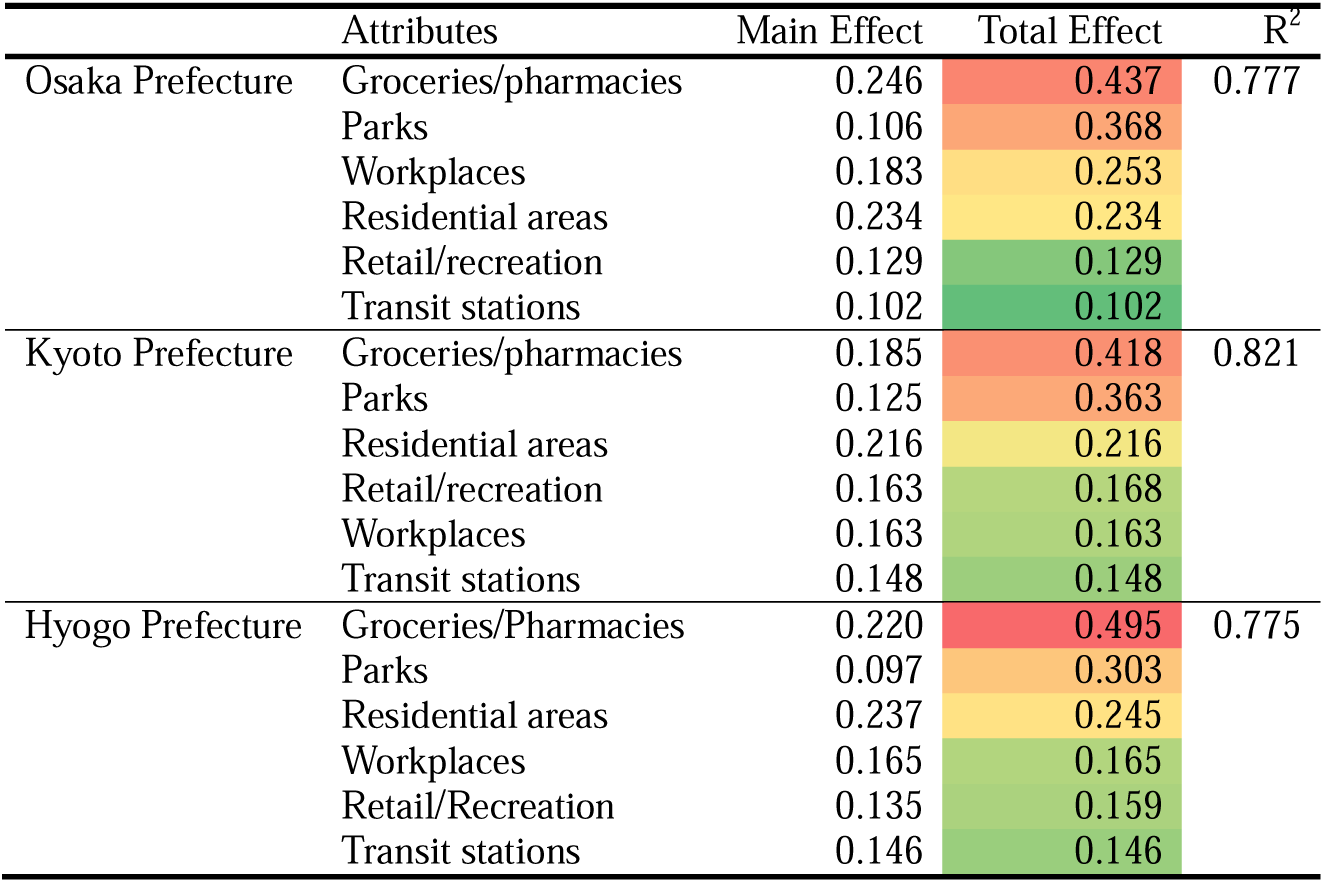
Main effects and total effects by prefecture. In Table 1, each number is represented by a green-to-red graduation. Specifically, the red tab has higher numbers, and the green tab has lower numbers.

**Figure 4.**
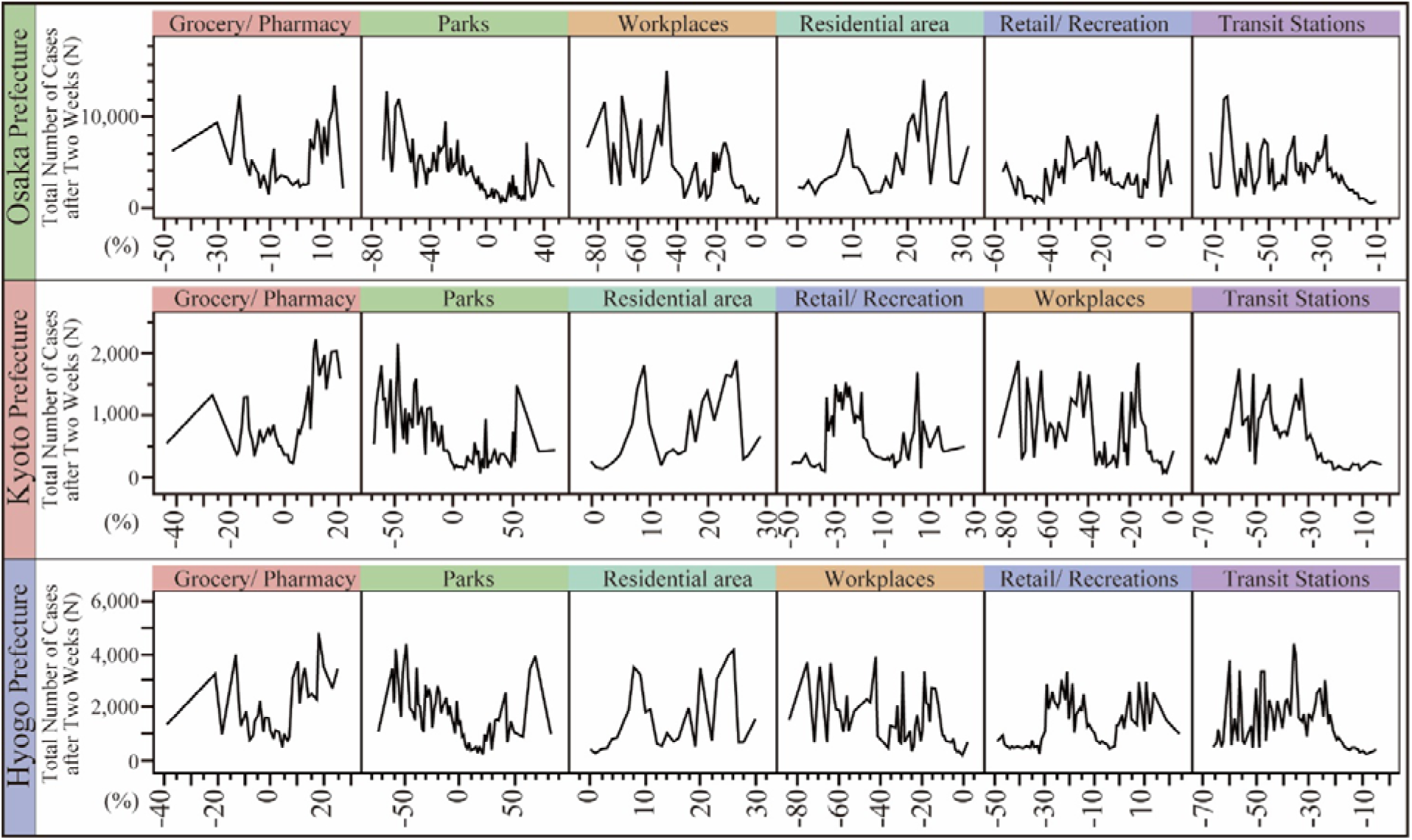
Variable importance by prefecture. The variable importance is assessed by the dependent resampled inputs, which are factor values constructed from observed combinations using a k-nearest neighbors approach.

In Osaka Prefecture, the R^2^ score was 0.777, which indicates that the model has good accuracy. The total effect was higher for human mobility in groceries/pharmacies (total effect=0.437), parks (total effect=0.368), workplaces (total effect=0.253), and residential areas (total effect=0.234). The total number of COVID-19 cases after two weeks gradually decreased by decreasing the human mobility of groceries/pharmacies by approximately 5% to -5%. In addition, the human mobility of parks increased from -20% to 20%, which reduced the number of COVID-19 cases. It was also found that the total effect was lower for human mobility in transit stations (total effect=0.102).

In Kyoto Prefecture, the R^2^ score was 0.821, which indicates that the model has high accuracy. The total effect was higher for human mobility in groceries/pharmacies (total effect=0.418), parks (total effect=0.363), and residential areas (total effect=0.216). The total number of COVID-19 cases after two weeks gradually decreased by decreasing the human mobility of groceries/pharmacies by approximately 10% to 5%. In addition, the human mobility of parks increased from - 20% to 50%, which reduced the number of COVID-19 cases. It was also found that the total effect was lower for human mobility in transit stations (total effect=0.148).

In Hyogo Prefecture, the R^2^ score was 0.775, which indicates that the model has good accuracy. The total effect was higher for human mobility in groceries/pharmacies (total effect=0.495), parks (total effect=0.303), and residential areas (total effect=0.245). The total number of COVID-19 cases after two weeks gradually decreased by decreasing the human mobility of groceries/pharmacies by approximately 10% to 0%. In addition, the human mobility of parks increased from - 50% to 10%, which reduced the number of COVID-19 cases. It was also found that the total effect was lower for human mobility in transit stations (total effect=0.146).

## 3. Discussion and Conclusion

### 3.1 Controlling Human Mobility

In conclusion, the results of this analysis indicate that it is essential to control the human mobility of groceries/pharmacies to less than 0% and the mobility of parks to more than -20%. This finding is important because human mobility control would reduce the number of people infected with SARS-CoV-2. To control the human mobility of groceries/pharmacies, the government must actively encourage residents to shop online and diversify the time spent using grocery stores and pharmacies. In fact, during the Delta variant outbreak in the Osaka metropolitan area, there were many incidents of infection clusters in groceries and department stores. Previously, the Japanese government did not restrict the shopping necessary to maintain daily life, even during the emergency declaration period. This study suggests that controlling the human mobility of groceries/pharmacies can prevent the rapid increase in the total number of cases after two weeks in the emergency declaration period. The results differ from those of a previous study [22]. This finding is significant because this study clarified the necessity of reducing the human mobility of grocery stores and pharmacies. The target value for reducing human mobility is under 0%.

The human mobility of parks was also found to impact the number of infections. As previous studies have found [21,22], increasing the human mobility of parks contributes to a decrease in the number of infections. This finding suggests that increasing the human mobility of parks decreases the number of infections. It means that parks could be actively used in the emergency declaration period instead of controlling human mobility in groceries/pharmacies.

The most significant finding for urban sustainability is that urban transit was not found to be a source of infection. The human mobility of transit stations has been used as a reference in policymaking [20]. Hence governments in cities around the world may be able to encourage communities to return to transit mobility if they can follow the kind of hygiene processes conducted in Osaka. Those hygiene processes are to maintain the number of daytime train departures even if the number of passengers decreases significantly [29]. Besides, effective hygiene processes are to provide incentives to those who take the train at times when the number of passengers is less. Those hygiene processes reduced the density of human mobility at the transit station [30]. The result suggest that governments could consider ending restrictions in transit stations where infections are less likely to occur. Increasing the number of people using public transportation would reduce air pollution such as carbon dioxide emissions.

This study also clarified that the government needs to consider the third most influential type of human mobility according to the characteristics of each prefecture. For example, the Osaka Prefecture government could reduce the number of infections by increasing the human mobility of workplaces to 0% by allowing people to go to work. The results suggest that it would be better for the government not to prevent people from going to work but rather to prevent them from shopping and other activities associated with work.

### 3.2 Urban Sustainability Implications

Currently, many people can be vaccinated against the disease in many countries. On the other hand, new variants of COVID-19 arise continuously. Therefore, it is unlikely that the pandemic will end, as the number of infections regularly increases and decreases. Human mobility control may continue to be the most effective method of a nonpharmaceutical intervention, even in the future. However, unlike in the early stages of the pandemic, it is not necessary to stop all human mobility through a lockdown. The results of this study indicate that the control of specific types of human mobility could have a greater effect according to the infection stage. For example, we could ask people to reduce their opportunities to go to groceries/pharmacies and inform people about the safety of going to parks and transit stations. This finding is important because it allows for the maintenance of social and economic activities even during pandemics for the post-COVID-19 pandemic.

However, the change might impact urban sustainability negatively during the medium-term COVID-19 pandemic. Air pollution was reduced by the human mobility limitation [34]. If governments simply reactivate human mobility, carbon dioxide emissions may increase due to car traffic [35]. Therefore, the government needs to conduct a mix of several policies, such as working from home, online shopping, and active use of public transportation. Those policies could contribute to improving the urban sustainability for the post COVID-19 pandemic.

The limitation of this study was that it was able to analyze only six types of human mobility available on Google Community Mobility Reports. Therefore, we cannot deny the possibility that the control of human mobility proposed by this study might cause an increase in another type of human mobility and a gradual increase in the number of infections. For example, would it truly be effective to restrict mainly dining and drinking establishments? To address this limitation, future research should research more diverse types of human mobility using GPS location history data. These GPS log data can be obtained at regular intervals from mobile phones with users’ consent. Using such data, we can clarify the relationship with the number of infections in more detail.

## 4. Methods

### 4.1. Human Mobility Data

This study used Google Community Mobility Reports data to analyze human mobility. The data are available publicly to provide insights into what has changed in response to policies aimed at combating COVID-19 [16]. The Google Community Mobility Reports chart movement trends by geography across six categories: retail/recreation, groceries/pharmacies, parks, transit stations, workplaces, and residential areas. Retail/recreation includes restaurants, cafes, shopping centers, theme parks, museums, libraries, and movie theaters. Groceries/pharmacies includes grocery stores, food warehouses, farmers markets, specialty food shops, drug stores, and pharmacies. Parks includes local parks, national parks, public beaches, marinas, dog parks, plazas, and public gardens. Transit stations includes public transport hubs, such as subway, bus, and train stations. The data show relative changes in visitors to the six types of places compared to the baseline days, which were the median value for the five weeks from January 3 to February 6, 2020 [36]. The data are published for each prefecture. This study uses data from the Osaka, Kyoto, and Hyogo Prefectures in the Osaka metropolitan area.

This research protocol was approved by the Research Ethics Committee of the Graduate School of Life Science, Osaka City University (No. 21-58). In addition, all methods used in this study followed the “Guidelines for the Use of Device Location Data,” which prohibit the use of GPS data for any purpose that involves identifying individual users to protect the privacy of users’ GPS location history [37].

### 4.2. Number of COVID-19 Cases Data

This study analyzed the daily number of newly confirmed cases of SARS-CoV-2 in the Osaka, Kyoto, and Hyogo Prefectures in the Osaka metropolitan area. The data were obtained from public information on COVID-19 infections provided by the Japanese Ministry of Health, Labour and Welfare [38]. The published data include the number of new COVID-19 cases each day by prefecture. The data do not include any personally identifiable information.

### 4.3 Statistical Analysis

The random forest method was used to analyze the relationship between the total number of COVID-19 cases after two weeks and the human mobility data. The random forest analysis of this study is an unsupervised analysis. Random forest predicts a response value by averaging the predicted response values across many decision trees [39]. Each tree is grown from a bootstrap sample of the training data. A bootstrap sample is a random sample of observations drawn with replacement. In addition, the predictors are sampled at each split in the decision tree [40]. Compared to machine learning, such as neural networks, random forests obtain highly accurate models with high R^2^ scores. For the statistical analysis, this study used JMP PRO 16.0.

The predictor variables are daily human mobility data of retail/recreation, groceries/pharmacies, parks, transit stations, workplaces, and residential areas. The response variable is the total number of COVID-19 cases after two weeks (fourteen days). The two-week lag is because SARS-CoV-2 takes approximately two weeks from infection to disease occurrence [41]. The effectiveness of Google mobility data was validated for 10-day forecasts of COVID-19 cases [42]. The number of trees in the forest is ten thousand for the random forest.

Based on the results of the random forest, this analysis focused on the R^2^ score, the main effect, the total effect, and the variable importance of the prediction profiler. The variable importance is assessed by the dependent resampled inputs, which are factor values constructed from observed combinations using a k-nearest neighbors approach. The assessed method is helpful if factors may be correlated with each other.

## Data Availability

The data presented in this study are available from references [16, 33].

## Funding

This research was funded by JSPS KAKENHI (grant number 21K14318) and the Dai-ichi Life Foundation (Incentive Research).

## Institutional Review Board Statement

The Research Ethics Committee of the Graduate School of Life Science, Osaka City University approved this research protocol (21-58).

## Informed Consent Statement

Informed consent was obtained from all subjects based on the privacy policy.

## Data Availability

The data presented in this study are available from references [16, 33].

## Conflicts of Interest

The authors declare no conflicts of interest.

## Notes

### Competing Interest Statement

The authors have declared no competing interest.

